# Estimating the size of undetected cases of the SARS-CoV-2 outbreak in Europe: An upper bound estimator

**DOI:** 10.1101/2020.07.14.20153445

**Authors:** Irene Rocchetti, Dankmar Böhning, Heinz Holling, Antonello Maruotti

**Affiliations:** Statistical Office - Consiglio Superiore della Magistratura; Southamption Statistical Sciences Research Institute, University of Southampton; Department of Methods and Statistics, Faculty of Psychology and Sports, University of Münster; Dipartimento di Giurisprudenza, Economia, Politica e Lingue Moderne, Libera Università Ss Maria Assunta, Department of Mathematics, University of Bergen

**Keywords:** Capture-recapture methods, COVID-19, Geometric distribution, Chao’s lower bound

## Abstract

**Background:** While the number of detected SARS-CoV-2 infections are widely available, an understanding of the extent of undetected cases is urgently needed for an effective tackling of the pandemic. The aim of this work is to estimate the true number of SARS-CoV-2 (detected and undetected) infections in several European Countries. The question being asked is: How many cases have actually occurred?

**Methods:** We propose an upper bound estimator under cumulative data distributions, in an open population, based on a day-wise estimator that allows for heterogeneity. The estimator is data-driven and can be easily computed from the distributions of daily cases and deaths. Uncertainty surrounding the estimates is obtained using bootstrap methods.

**Results:** We focus on the ratio of the total estimated cases to the observed cases at April 17th. Differences arise at the Country level, and we get estimates ranging from the 3.93 times of Norway to the 7.94 times of France. Accurate estimates are obtained, as bootstrap-based intervals are rather narrow.

**Conclusions:** Many parametric or semi-parametric models have been developed to estimate the population size from aggregated counts leading to an approximation of the missed population and/or to the estimate of the threshold under which the number of missed people cannot fall (i.e. a lower bound). Here, we provide a methodological contribution introducing an upper bound estimator and provide reliable estimates on the *dark number*, i.e. how many undetected cases are going around for several European Countries, where the epidemic spreads differently.

## 1 Introduction

The severe acute respiratory syndrome coronavirus 2 (SARS-CoV-2) has become a pandemic within few weeks. The number of detected cases increased day-by-day, at an exponential rate at the beginning, and now follows a logistic distribution [1, 2]. Cases of SARS-CoV-2 might have been vastly under-reported in official statistics. It is widely acknowledged that the majority of the cases are asymptomatic and, thus, not observed or recorded [3–5]. In other words, the available data just tell us a part of the story: individuals may be already infected but are not aware of it, maybe because of the absence of symptoms, or cases may be under symptomatic suspicion but the disease has not been diagnosed yet (due to the delay in getting swab results). The total number of cases is thus unknown, and general comments on the spread of the epidemic are thus partial as based on a (relatively small) fraction of the total cases. Some studies have used simulation-based approaches to infer reasonable estimates of total number of cases, but often these estimates are surrounded by poor uncertainty measures, leading to too wide confidence intervals [6]. Here, we are proposing a simple and effective method to obtain reasonable point and interval estimates of the total number of SARS-CoV-2 infections in several European countries. In detail, we introduce a novel estimator based on a capture recapture (CR) approach. The capture-recapture method should be considered the gold standard for counting when it is impossible to identify each case and large undercounts will occur [7]. CR methods were originally developed in the ecological setting with the aim of estimating the unknown size of a (possibly elusive) population and then they started to be applied also to epidemiological and health sectors (see [8, 9]). Many CR estimators have been proposed in the literature (see e.g. [10–12]), and some of them can be used to identify lower bounds [13] of the population size. In the analysis SARS-CoV-2 infections, official data are available at the aggregated level, whereas individual data are not available to the general or the academic public. Hence, it is not possible to get the exact distribution of the number of infected individuals observed exactly one day, exactly two days and so on until *m* days. The population is open, subjected to deaths, and this may further complicate the analysis [14]. A lower bound of the total number of infected cases is computed by [3] modifying the Chao estimator [15] to address issues related to the data at hand. This is a relevant result as it provides reasonable information to the policy makers about the undetected cases and the magnitude this phenomenon may have at least, so that national health systems may be aware of the minimum number of cases that may demand health care services. At this stage of the spread of the epidemic, governments are willing to relax restrictive measures and several researches address issues related to the epidemic [16–18]. To calibrate the new interventions, an estimate of the lower bound of the number of infections may not be enough, as SARS-CoV-2 has already shown to spread around the population very quickly [19–21]. This contribution aims at providing an approximated upper bound for the total number of SARS-CoV-2 cases, to better appreciate the dimension of the epidemic, under the worse scenario. Such an estimate is obtained from a non parametric CR model, providing an upper bound estimate of the total number of infections regardless of the true data generating process.

This contribution is organized as follows. In Section 2, we introduce the basic notation and how we are going to work with the data at hand. A brief summary of the modified Chao lower bound is also discussed. These notions are then used to compute the upper bound, details of which are provided in Section 3, along with the computation of the uncertainty surrounding the estimates. In Section 4 we show the empirical application of the proposal on data from several European countries. A discussion showing other interesting insights concludes.

## 2 Methods

### 2.1 Preliminaries

Let us denote with *N* (*t*) the cumulative count of infections at day *t* where *t* = *t*_0_, *…, t*_*m*_. Hence Δ*N* (*t*) = *N* (*t*) *− N* (*t −* 1) are the number of new infections at day *t* where *t* = *t*_0_ + 1, *…, t*_*m*_. Also, let *D*(*t*) denote the cumulative count of deaths at day *t* where *t* = *t*_0_, *…, t*_*m*_. *t*_0_ defines the beginning of the observational period and *t*_*m*_ defines the end. We assume the trivial assumption *t*_*m*_ *> t*_0_, so that the observational window is not empty. Again, we denote with Δ*D*(*t*) = *D*(*t*) *− D*(*t −* 1) the count of new deaths at day *t* where *t* = *t*_0_ + 1, *…, t*_*m*_.

The question arises how this can be linked to a capture-recapture approach. Let *X*_*i*_ denote the number of identifications for each infected individual *i* typically provided by the days the individual will surely remain infected. Let denote *τ*_*x*_ the probability of identifying an individual *x* times where *x* = 0, … A lower bound estimator of the unobserved frequency *f*_0_, say 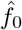, can be estimated by using the observed frequency of those identified exactly once, *f*_1_, and of those identified twice, *f*_2_, [13, 15]:

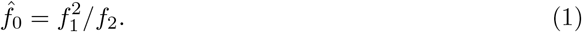

It is thus crucial to relate *f*_1_ and *f*_2_ with the data at hand. In detail, at each day *t, f*_1_(*t*) represents the infected people identified just once, i.e. the new infections, whose number is given by Δ*N* (*t*). Similarly, *f*_2_(*t*) represents the infected people detected at time (*t −* 1) and still infected at time *t*. This can be computed as Δ*N* (*t −* 1) *−* Δ*D*(*t*). Hence the estimate for the number of hidden infections at day *t* is

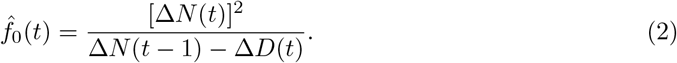

By applying the estimator (1) day-wise we get the modified Chao lower bound estimator (see [3]):

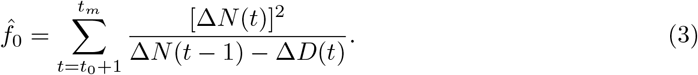

In practice, however, the bias-corrected form of (3) suggested by [22] is used:

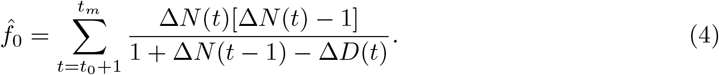

We define the understanding that Δ*N* (*t −* 1) *−* Δ*D*(*t*) is set to 0 if it becomes negative, in other words we use *max*{0, Δ*N* (*t −* 1) *−* Δ*D*(*t*)}. The final estimate of lower bound (LB) of the total number of infection is then given as what has been observed at the end of the observational window *t*_*m*_ and the estimate of the hidden numbers:

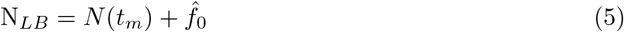

### 2.2 The Upper Bound estimator

The lower bound is helpful as an indication of the minimum number of people having had SARS-CoV-2 and answers to a fundamental open question: “How many undetected cases are at least going around?”. Nevertheless, this information may be treated as a starting point whenever interventions and tools to dampen the spread of the epidemic are rolled out. The proposed upper bound estimator extends the research on the undetected cases and helps policy makers to evaluate the SARS-CoV-2 epidemic situation locally and at the current phase of its development. An estimate of the worse possible scenario is provided.

Following a similar strategy as in Section 2.1, this is achieved by firstly estimating daily-specific upper-bounds and then summing up all the estimates to get the final point-estimate of the maximum number of undetected cases. This daily–wise based upper bound approach provides an approximation of the data generation process.

Let us introduce the cumulative distribution function

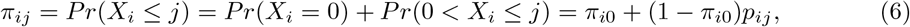

where homogeneity in the probability of being infected at a certain date *t* is assumed, i.e. *π*_*ij*_ = *π*_*j*_, with *p*_*ij*_ = *p*_*j*_ being the cumulative zero-truncated probability distribution. Equation (6) represents the probability that an individual is infected for at most *j* days, and it is function of *π*_0_ and *p*_*j*_; but *π*_0_ is not observed. The quantities *p*_*j*_(*j* = 1, 2, 3) in equation (6) at each time *t* may be approximated as

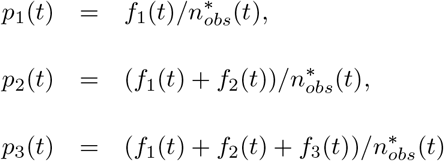

where *f*_1_(*t*) and *f*_2_(*t*) have been introduced in the previous section and

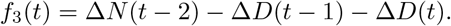

and 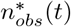 is the number of current infected individuals observed at each time. We think that it is reasonable, for each day *t*, to consider the number of individuals affected by SARS-CoV-2 for the day *t*, for day *t* and the day before, and, for day *t* and the two days before, as *m* = 3 is the minimum number of consecutive days of new infections necessary for the upper bound estimator to be computed. Furthermore, considering more than 3 days for an individual to be observed as affected by SARS-CoV-2 would lead to the risk of not observing the number of people affected by SARS-CoV-2 for exactly four, five and so on times because of the higher risk of overlapping cases.

Since *π*_0_ is unknown, to compute the probabilities in (6), we substitute it with

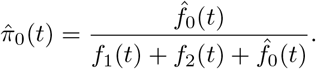

where 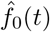 is the *lower bound* probability of undetected cases derived from the Chao estimator in its bias corrected form, computed at each time *t* (see Equation 2). This also explains why a lot of detail was devoted to the lower bound estimator in the previous section as it is very much needed here. In other words, based on the Chao lower bound estimator of the undetected cases, we derive the *complete* count distribution and calculate the upper bound for the population size on such a complete distribution. Now, it follows that Equation (6) takes the form

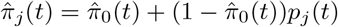

when theoretical probabilities are replaced by their now available estimates. In order to provide an upper bound estimator we use the main results of [23]:

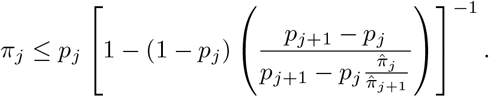

For *j* = *m −* 2, and by some algebra, we get the equivalent condition

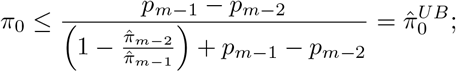

that makes clear why at least *m* = 3 days should be considered. The right-hand side 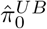 of the above inequality provides an upper bound estimate of the population size based on the Horvitz–Thompson estimator:

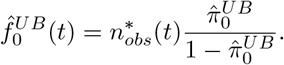

However we deal with a day-wise upper bound approximation of *π*_0_(t) which is given by

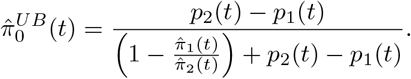

To get an estimate for the missed SARS-CoV-2 infections 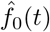 at each time *t* we compute the Horvitz–Thompson (HT) estimator at each time *t* and ultimately we sum it up over all times, reaching thus the final upper bound for the missed SARS-CoV-2 cases *n*_0_ as follows

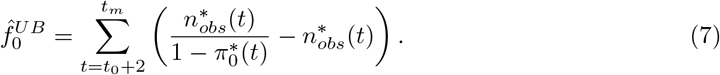

Hence, the approximated upper bound of the total number of infected people, 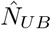, in the time window from *t*_0_ to *t*_*m*_ is then given by

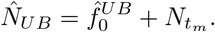

### 2.3 Uncertainty estimation

A fundamental issue in general CR analyses is the quantification of uncertainty surrounding the estimates of the unknown population size. An estimation of the population size can be correctly computed, but if the associated estimation of variance is poor, then coverage by the 95% confidence interval may falsely indicate poor estimation by the point estimator, i.e. the point estimator may result in a poor coverage rate. Focusing on the proposed upper-bound estimator, we attempt here to investigate bootstrap methods as a robust and general approach to estimate variances and confidence intervals. Various bootstrap methods have been considered to estimate uncertainty in CR analyses with respect to other estimators [24–27]. In the following, we consider two different bootstrap approaches to approximate the uncertainty surrounding the point estimate: the imputed and the reduced bootstrap approaches.

Under the imputed bootstrap approach, we draw 1000 bootstrapped samples of size *N*_*UB*_ generated according to a multinomial model whose probabilities are given by

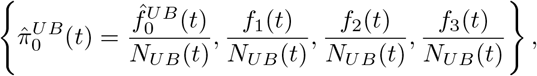

where 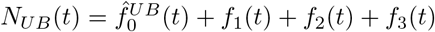.

Differently, under the reduced bootstrap approach, each of the bootstrapped samples contains 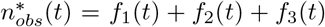 observations generated according to a multinomial model whose probabilities are given by 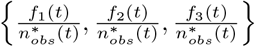. For each of the two approaches, the upper bound *N*_*UB*_ is computed for each bootstrapped sample, by summing up over the time period. Of course, for the imputed bootstrap the fraction of undetected cases is dropped and considered unknown when computing the population size. We report the 2.5% and 97.5% values of *N*_*UB*_ distribution. This allows us to overcome issues often encountered in the construction of the symmetric confidence intervals [28]: the sampling distribution could be skewed, the coverage probabilities may be unsatisfactory, etc.

## 3 Data Analysis

The example provided here relies on European data. The time series of cumulative cases and deaths up to 17/04/2020 are considered and are taken from https://github.com/open-covid-19/data. A graphical representation of the data at hand is shown in Figure 1.

**Figure 1:**
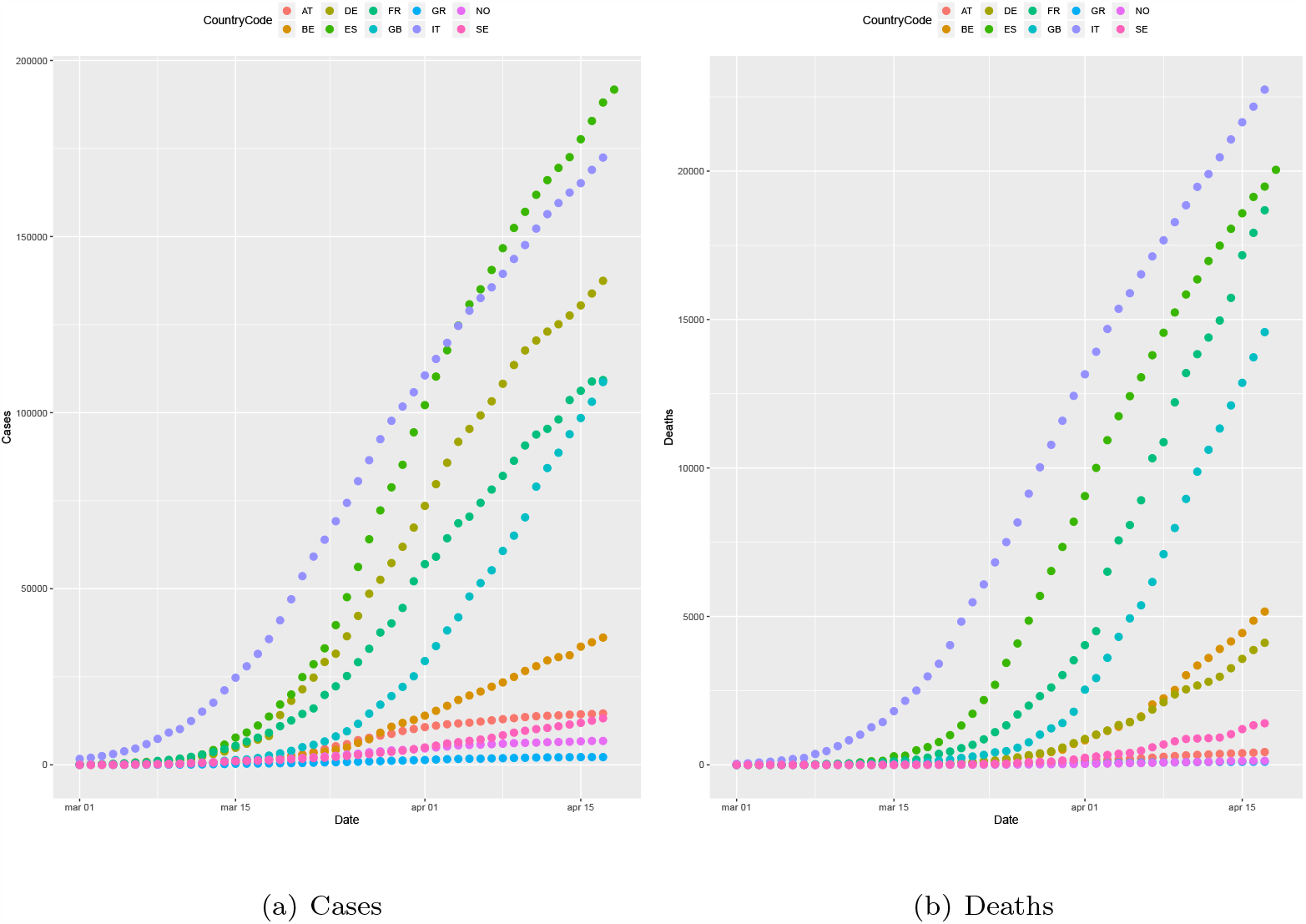
Cases and deaths for the analyzed countries.

Data from the day which we record the first death are analyzed only. We obtain the estimates of an upper bound for undetected cases for several European countries (see Table 1). The last column in Table 1 shows the ratio of the total estimated cases to the observed cases. The ratio of the total estimated cases (in the worse scenario) to the observed cases is interesting in itself. A ratio of 4.5 would mean that for every observed patient there are 3.5 infected persons unseen. The reason for this can be manifold as these unseen cases might be without symptoms or show very mild signs of infection.

**Table 1:**
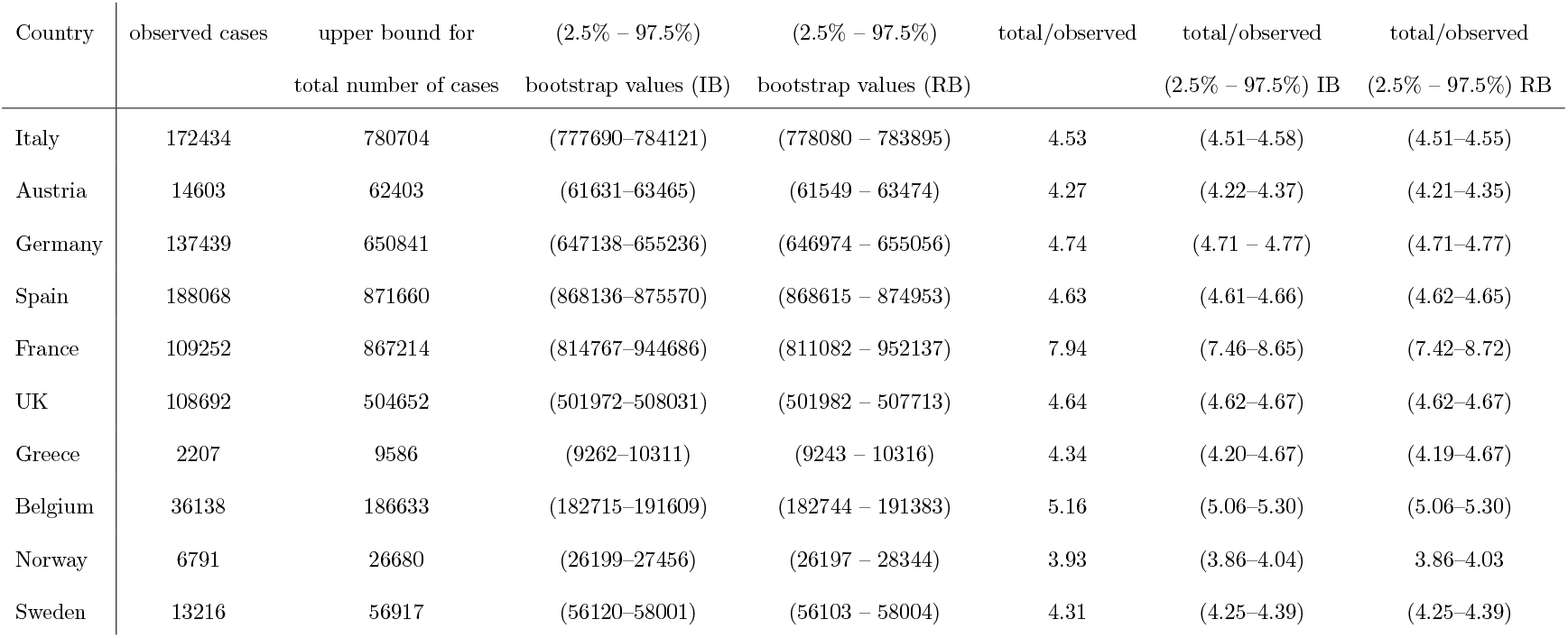
Estimated hidden and total cases of Sars-Cov-2 for several European countries, at 17/04/2020.

As expected, the undetected cases represent a relevant portion of the total number of cases. This is in line with a few existing works and discussions on the topic, see e.g. [29–31]. The number of total number of cases are at most approximately 4.5 times the observed cases. Of course, differences arise at the Country level, and heterogeneous estimates ranging from the 3.93 times of Norway to the 7.94 times of France, see Table 1. These differences are due to different heterogeneity structures in the cases and deaths time series at the country level. These results are telling us that SARS-COV-2 outbreak was more prevalent than described by the official data, though a significant number of individuals that are infected actually remain asymptomatic.

Point estimates can be used to synthetically describe the SARS-COV-2 outbreak, but they may be rather uncertain. In Table 1, we also provide uncertainty measures, based on the boot-strap procedures described in Section 3.1. It is also possible to compare the two employed bootstrap approaches. They perform rather similarly (see also [26]) and the bootstrap intervals are rather narrow, with France only showing a rather wide interval to indicate that its point estimate should taken with caution.

## 4 Conclusions

Different capture-recapture approaches have been used to estimate the size of a partially observed population; many parametric or semi-parametric models have been developed to estimate the population size from aggregated counts leading to an approximation of the missed population and/or to the estimate of the threshold under which the number of missed people cannot fall (i.e. a lower bound). While several proposals for the latter exist, the estimation of an upper bound in capture recapture methods has been often overlooked, with the exception of the recent work of [23]. We propose an extension of the upper bound estimator under cumulative data distributions, in an open population, such that a day-wise estimator varying over time. The approach results in a time-aggregated approximation for *f*_0_ and thus for *N*. The proposed upper bound estimator has been applied to registered cases in some European Countries; confidence intervals for *N* have been provided by employing bootstrap approaches. We consider, for each country, data up to the 17 of April, by assuming, given also the day wise nature of the estimator, that the recoveries are negligible; however when dealing with cases and deaths at a more recent date, given the increased percentage of immune people, recoveries should be taken into account in the computation.Another issue which should be considered is the one concerning the role of the deaths: even when the number of confirmed cases for two different Countries are close to each other the upper bounds can be different according to the deaths size wrt the cases (i.e. France and Spain). The length of the observation window plays an important role in this context and according to the distribution of SARS-CoV-2 cases observed more than once, the distribution can be less or more stable. It appears necessary to analyze this issue more deeply and we propose to do this in a future work.

## Data Availability

All data used in this manuscript are publicly available.

